# A spectrum of recessiveness among Mendelian disease variants in UK Biobank

**DOI:** 10.1101/2021.12.13.21267756

**Authors:** Alison R. Barton, Margaux L.A. Hujoel, Ronen E. Mukamel, Maxwell A. Sherman, Po-Ru Loh

## Abstract

Recent work has found increasing evidence of mitigated, incompletely penetrant phenotypes in heterozygous carriers of recessive Mendelian disease variants. We leveraged whole-exome imputation within the full UK Biobank cohort (*N*∼500K) to extend such analyses to 3,481 rare variants curated from ClinVar and OMIM. Testing these variants for association with 57 quantitative traits yielded 103 significant associations involving variants previously implicated in 35 different diseases. Notable examples included a *POR* missense variant implicated in Antley-Bixler syndrome that associated with a 1.76 (s.e. 0.27) cm increase in height, and an *ABCA3* missense variant implicated in interstitial lung disease that associated with reduced FEV1/FVC ratio. Association analyses with 1,257 disease traits yielded five additional variant-disease associations. We also observed contrasting levels of recessiveness between two more-common, classical Mendelian diseases. Carriers of cystic fibrosis variants exhibited increased risk of several mitigated disease phenotypes, whereas carriers of spinal muscular atrophy alleles showed no evidence of altered phenotypes. Incomplete penetrance of cystic fibrosis carrier phenotypes did not appear to be mediated by common allelic variation on the functional haplotype. Our results show that many disease-associated recessive variants can produce mitigated phenotypes in heterozygous carriers and motivate further work exploring penetrance mechanisms.

## Introduction

Since the advent of next generation sequencing, the number of variants identified as contributing to Mendelian disease has grown rapidly^1^. Roughly 20% of all protein-coding genes in humans have been associated with at least one Mendelian disease^2^. Increasingly, studies of recessive disease variants have begun observing that these variants can sometimes cause mitigated phenotypes in heterozygous carriers, thereby contributing to population variation in complex traits and disease susceptibility^3–11^. However, the rarity of most such variants together with their unavailability in most SNP-array-based genotyping studies has limited attempts to explore this phenomenon at scale. Early work focused on smaller cohorts recruited for specific diseases, such as a series of studies that demonstrated increased risk of male infertility^12,13^, bronchiectasis^14–16^, and asthma^15,17^ among other phenotypes in cystic fibrosis carriers. More recently, larger data sets have enabled extending the breadth of such analyses to more phenotypes^4,5^ and to more recessive disease variants^3^.

With increasing exome sequencing of population biobank cohorts^18–20^, a new opportunity to search for carrier phenotypes in a phenome-wide, exome-wide manner has emerged. Furthermore, biobank data sets present an opportunity to ameliorate ascertainment biases by assessing phenotypes in population cohorts, complementing analyses of patients and their families. Family-based studies have been observed to be susceptible to ascertainment biases that inflate observed effects^21^, while the opposite “healthy volunteer” phenomenon has been observed in biobank cohorts^22^. Genome-wide genotyping and imputation in biobank data sets also provide opportunities to investigate potential genetic modifiers of incompletely penetrant carrier phenotypes^23^.

Here we leveraged exome-wide imputation within the UK Biobank cohort^24^ to power a broad investigation of quantitative and disease phenotypes amongst carriers of recessive disease variants. Next, we performed a focused analysis of two relatively more common severe recessive Mendelian diseases, using the power afforded by high carrier frequencies to characterize carrier phenotypes or establish a truly recessive pattern of phenotypes. Finally, we considered the molecular mechanisms underlying incomplete penetrance observed amongst carriers, evaluating a previously proposed model of modified penetrance^23^.

## Subjects and methods

### Imputed carriers of recessive Mendelian disease variants in UK Biobank

We previously used the first tranche of whole exome sequencing data released by the UK Biobank (*N*=49,960)^18^ to impute coding variants into SNP-array data available for *N=*487,409 participants in the full UK Biobank cohort^25^, achieving accurate imputation of rare variant genotypes at minor allele frequencies (MAF) down to ∼0.00005^24^. Here, we analyzed a subset of imputed variants that were annotated in ClinVar^26^ as “pathogenic” or “likely pathogenic” for diseases annotated in OMIM^2^ as “autosomal recessive”. We further restricted to rare variants (MAF < 0.01) with a minimum MAF of 0.00001 and estimated imputation accuracy of *R*^2^ > 0.5, leaving 3,481 variants for analysis.

### Association tests with quantitative traits

We tested imputed genotype dosages for association with 57 quantitative traits using linear mixed models implemented in BOLT-LMM v2.3.4. These traits included the 54 quantitative traits we previously analyzed^24^ and three additional traits (skin pigmentation, tanning ability, and hair color). We performed quantitative trait association analyses on *N*=459,327 UK Biobank participants who reported European ancestry and had not withdrawn from the study at the time of analysis. We did not attempt to filter homozygotes or compound heterozygotes from these analyses, reasoning that such individuals would account for negligible numbers of carriers of the rare variants we analyzed (both based on allele frequencies and on the “healthy volunteer” ascertainment bias of UK Biobank).

### Association tests with binary traits

We tested the same imputed variants for association with 1,139 binary disease phenotypes curated by UK Biobank. These consisted of the complete set of “first-occurrence” of disease traits in the UK Biobank converted to simple case and control status as well as the set of 8 “algorithmically defined health outcomes” disease categories provided by the UK Biobank. We tested variants for association with binary traits using the BinomiRare test^27^ to obtain *p*-values robust to case-control imbalance while adjusting for age (stratified into five-year tranches) and sex. For computational efficiency, we re-implemented the BinomiRare test and applied a binomial approximation when the number of observed cases among carriers exceeded 100. We estimated odds ratios as *xw* / *yz*, where *x, y, z, w* denote ratios of observed versus expected cases among carriers, cases among noncarriers, controls among carriers, and controls among noncarriers, respectively. We estimated 95% confidence intervals using a normal approximation, i.e., converting *p*-values to *z*-scores and then taking the 95% CI of the log odds ratio (OR) to be log(OR) ± 1.96 * logOR / *z*. We performed association analyses on an unrelated subset of *N*=415,291 UK Biobank participants who reported European ancestry and had not withdrawn from the study^28^.

### Analyses of cystic fibrosis carriers

We identified cystic fibrosis carriers in UK Biobank using SNP-array genotypes for the Phe508del variant and (in auxiliary analyses) the missense SNP rs78655421, excluding participants with a cystic fibrosis report (according to the “first occurrences” data field). We applied the same analysis pipeline as above to test for associations with the 1,139 binary traits and applied a significance threshold of FDR < 5% (*q*-value < 0.05).

### Analyses of spinal muscular atrophy carriers

We identified spinal muscular atrophy carriers in the UK Biobank *N*=200K exome sequencing release as individuals with evidence of only one functional copy of *SMN1*. We estimated the number of functional copies of each of *SMN1* and *SMN2* based on depth of coverage of exome sequencing reads that mapped uniquely to the exon 7-intron 7 region of each gene (chr5:70,951,800-70,952,600 for *SMN1* and chr5:70,076,400-70,077,100 for *SMN2* in hg38 coordinates; these regions contain four paralogous sequence variants that distinguish the highly homologous genes and were captured by exome sequencing). This approach accounted for deletions of exons 7-8 that commonly inactivate copies of *SMN2* and occasionally *SMN1*^29^. We computed exome sequencing read-depth using mosdepth v0.3.1^30^ and normalized each sample’s read-depth measurements against corresponding measurements from other samples with similar exome-wide sequencing depth profiles using a pipeline we recently described^28^.

We analyzed SMA carriers for evidence of changes in three traits related to neuromuscular function: walking speed, hand grip strength (maximum of left- and right-hand measurements), and FEV1 / FVC ratio (a measure of lung function). Using age, age squared, and sex as covariates, we performed linear regressions to test for an association between SMA carrier status and each trait.

### Testing a model of modified penetrance in carriers of loss-of-function variants

To further investigate potential molecular mechanisms underlying why some recessive variant carriers display mild phenotypes but others do not, we considered a model of modified penetrance proposed by Castel *et al*. (2018)^23^. This model proposes that the penetrance of a deleterious variant can be affected by variants on the allele on the homologous chromosome, particularly in the case of common *cis*-eQTLs that modulate expression of the functional copy of the gene. To evaluate this model, we analyzed heterozygous carriers of relatively common disease variants in two genes, *FLG* and *CFTR*. To perform association tests on variants carried on the haplotypes opposite the disease variants, we imputed variants from the Haplotype Reference Consortium panel (r1.1) using Minimac3 v2.0.1 (run on genomic windows including 3 Mb flanks of each gene) and analyzed these variants together with the variants we previously imputed from whole-exome sequencing^24^. Next, we extracted the genotypes for these carriers at variants within 1 Mb up- and downstream of each gene. We then recoded the genotypes for each carrier to be hemizygous for the alleles sitting on the haplotype opposite from the deleterious variant. We performed association tests on these recoded hemizygous variants using the Fisher’s exact test implemented in plink (v1.9)^31^ (--fisher-midp) (which could perform this analysis after we recoded the chromosome as “X” and coded all individuals as male). We assessed the power of these analyses to detect associations between common variants on the opposite haplotype using the wp.logistic function in the WebPower R package.

## Results

### Quantitative phenotypes in carriers of recessive disease variants

Testing 3,481 rare recessive disease variants for association with 57 quantitative traits measured in the UK Biobank identified 103 significant (*p* < 2.52 × 10^−7^; Bonferroni-corrected) variant-trait associations (**Fig. 1** and **Table S1**). These associations involved variants reported to be pathogenic for 35 distinct recessive diseases. For many of these diseases (19/35 diseases), carriers exhibited significant deviations in multiple quantitative traits. Some of these multiple associations partly reflected correlated measurements of blood, lipid, or pigmentation traits, such as associations between a variant believed to cause Bernard-Soulier syndrome type C (a recessive bleeding disorder) and mean platelet volume (0.68±0.03 SD), platelet distribution width (0.54±0.03 SD), and platelet count (−0.65±0.03 SD). However, others pointed to distinct manifestations of pleiotropy such as associations of a variant for McArdle disease (a recessive glycogen storage disorder that interferes with muscle function) with both increased urate levels (0.15±0.02 SD) and increased waist-hip ratio (0.10±0.02 SD).

**Figure 1.**
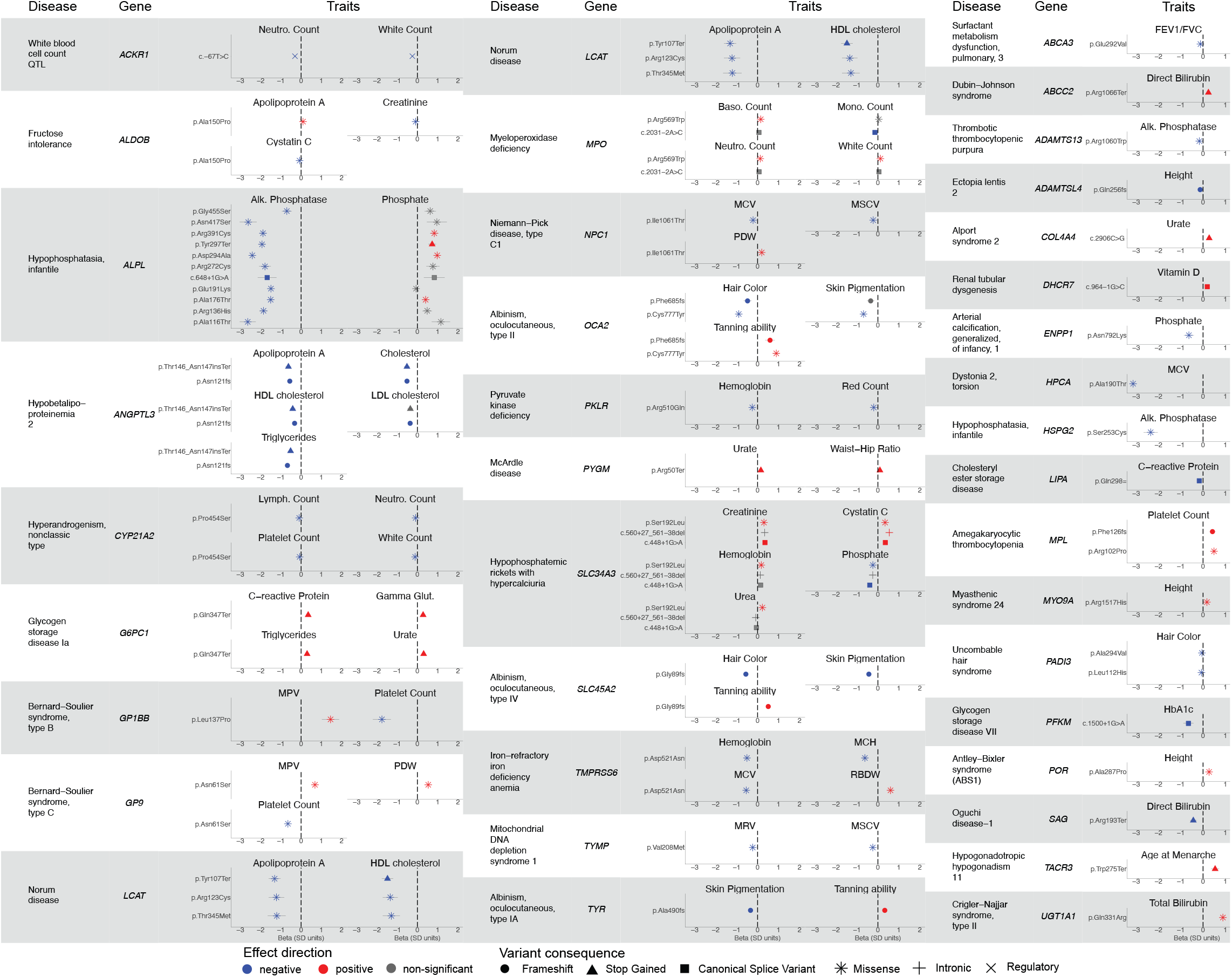
Carriers of recessive Mendelian disease variants display quantitative phenotypes. Mendelian diseases and their associated genes are listed to the left in each column, and the mean effect size is plotted on the right for each associated quantitative trait in units of standard deviation (error bars, 95% CIs). Positive-effect variants are shown in red, negative-effect variants in blue, and variants not Bonferroni-significant for one of the displayed traits in gray. Marker shapes correspond to effects on the gene and gene product as reported in ClinVar.

For some of the diseases, carriers exhibited traits that might be expected based on the known biological mechanisms of the disease, supporting the validity of our analytical approach. For example, when considering Mendelian disorders where the production of a particular protein or compound is altered, one might expect a carrier to have reduced levels of that same molecule. We observed this phenomenon with infantile hypophosphatasia, which is defined by errors in alkaline phosphatase^32^. In UK Biobank, carriers of several variants in the *ALPL* gene reported as pathogenic for recessive hypophosphatasia exhibited decreased alkaline phosphatase (ranging from -2.63±0.20 SD to - 0.71±0.12 SD) and increased phosphate, as might be expected. Another example involved two variants in *ANGPTL3* that have been implicated in hypobetalipoproteinemia 2, a recessive disorder in which individuals experience low levels of several lipid biomarkers^33^. Carriers showed decreases in apolipoprotein A levels (−0.56±0.04 SD; -0.63±0.07 SD), cholesterol levels (−0.52±0.04 SD; - 0.52±0.07 SD), and triglyceride levels (−0.67±0.04 SD; -0.52±0.07 SD).

Other diseases with more complex biological mechanisms yielded less straightforward carrier phenotypes. Here we highlight three such examples. First, a missense variant in *POR* implicated in Antley-Bixler syndrome^34^, a recessive skeletal disorder in which bones fuse prematurely, associated with a 0.27±0.04 SD increase in height (i.e., 1.76±0.27 cm). Second, a frameshift variant in *ADAMTSL4* implicated in ectopia lentis 2, a recessive disorder of the fibers in the eyes that can lead to vision problems, associated with a 0.13±0.02 SD (0.84±0.12 cm) decrease in height. Decreased height has been observed in patients with ectopia lentis 2, but the mechanism by which *ADAMTSL4* causes this change has not been extensively examined^35^. Third, a missense variant in *ABCA3* implicated in pulmonary surfactant metabolism dysfunction 3, a recessive interstitial lung disease caused by disruptions in the surface tension of lung surfactant^36^, associated with a 0.12±0.01 SD decrease in FEV1/FVC ratio, a measure of lung function. These examples add to the growing body of evidence that rare variants that cause severe disease in homozygotes or compound heterozygotes can often produce mild, subclinical phenotypes in heterozygous carriers^3,4^.

### Disease phenotypes in heterozygous carriers of recessive variants

We next tested the same set of 3,481 rare recessive disease variants for association with 1,139 binary traits in UK Biobank, identifying five associations that reached significance (*p* < 1.26 × 10^−8^; Bonferroni-corrected) (**Table 1**). As with the quantitative traits, some associations were expected from previous literature. Carriers of a frameshift variant in *HBB* exhibited increased risk of thalassemia^37^, and carriers of a stop gain variant in *COL4A4* implicated in Alport syndrome 2, a recessive disorder that involves kidney dysfunction, exhibited increased risk of hematuria (OR=10.5; 95% CI, 5.2-21.2)^38,39^, as we and others have recently reported^24,40,41^. Carriers of a missense variant in *TYR* (tyrosinase) implicated in recessive oculocutaneous albinism type IA exhibited increased risk of disorders of aromatic amino-acid metabolism (OR=63.3; 95% CI, 16.3-245.1)^42^. A missense variant in *TG* (thyroglobulin) implicated in recessive thyroid dyshormonogenesis 3 increased risk of hypothyroidism in carriers (OR=2.20; 95% CI, 1.68-2.88)^43,44^.

**Table 1.** Carriers of Mendelian recessive disease variants exhibit increased risk of less-severe disease phenotypes. Odds ratios and p-values are reported for the five associations that reached Bonferroni significance.

| Recessive disease association | ClinVar reported variant |  |  |  | Disease association in carriers | Trait category | OR (95% CI) | P-Value |
| --- | --- | --- | --- | --- | --- | --- | --- | --- |
|  | Gene | Variant | Variant impact | MAF |  |  |  |  |
| Alport syndrome 2 | COL4A4 | 2:227917083:G:C | stop gained | 6.16E-04 | Recurrent and persistent haematuria (N02) | Genitourinary system disorders | 10.47 (5.17-21.2) | 6.75E-11 |
| Thyroid dyshormonogenesis 3 | TG | 8:133894854:C:T | stop gained | 6.38E-04 | Other hypothyroidism (E03) | Endocrine, nutritional and metabolic diseases | 2.20 (1.68-2.88) | 9.72E-09 |
| Sickle cell anemia | HBB | 11:5248233:CAG:C | frameshift | 2.15E-05 | Thalassaemia (D56) | Blood, blood-forming organs and certain immune disorders | 3,183 (440-23,030) | 1.36E-15 |
| Albinism, oculo-cutaneous, type 1A | TYR | 11:88961072:C:A | missense | 1.10E-03 | Disorders of aromatic amino-acid metabolism (E70) | Endocrine, nutritional and metabolic diseases | 63.27 (16.33-245.10) | 1.94E-09 |
| Retinitis pigmentosa 80 & short rib thoracic dysplasia 9 | IFT140 | 16:1607935:C:A | splice donor | 5.11E-04 | Cystic kidney disease (Q61) | Congenital disruptions and chromosomal abnormalities | 18.42 (8.47-40.05) | 1.98E-13 |

A more intriguing association involved a splice donor variant in *IFT140* previously implicated in recessive short-rib thoracic dysplasia 9 and retinitis pigmentosa 80, often with accompanying renal disease^45,46^. Carriers of this variant exhibited increased risk of cystic kidney disease (OR=18.4; 95% CI, 8.5-40.1), corroborating recent findings from analyses of directly-sequenced individuals and imputation using the TOPMed reference panel^47,48^. Loss-of-function of both copies of *IFT140* appears to be inviable based on murine studies^49^, such that this canonical splice variant has been observed in cases of recessive disease only in compound heterozygotes with partial function of the other copy of the gene^50^. While retinitis pigmentosa 80 primarily manifests in visual symptoms and recessive short-rib thoracic dysplasia 9 in skeletal symptoms, *IFT140* encodes a protein related to cilia function that also is expressed in the kidney, and renal symptoms have been noted in both diseases. The observed association between carriers of the splice variant and cystic kidney disease suggests partial haploinsufficiency of *IFT140* in its role in the kidney.

### Contrasting recessiveness of cystic fibrosis and spinal muscular atrophy

In light of the diversity of autosomal recessive Mendelian diseases for which we observed mitigated phenotypes in carriers, we decided to more closely investigate two relatively common recessive Mendelian diseases to ask whether mitigated phenotypes were a ubiquitous feature of recessive disease carriers. To do so, we identified diseases with sufficiently high carrier frequencies in UK Biobank that we would be well-powered to identify mitigated carrier phenotypes or lack thereof. The two diseases we identified based on these criteria were cystic fibrosis (CF) and spinal muscular atrophy (SMA).

Previous studies have identified mitigated phenotypes in carriers of cystic fibrosis variants related to phenotypic manifestations of the disease^4,5^. To further explore the extent of this phenomenon utilizing the deep phenotyping of UK Biobank, we tested our full set of quantitative and binary traits for associations with carriers of the most common CF mutation, *CFTR* Phe508del (MAF=1.6%), which was directly genotyped by UK Biobank SNP-arrays. Carriers of this variant showed significant associations (*q*-value < 0.05) with asthma (OR=1.12; 95% CI,1.06-1.17), aspergillosis (OR=2.60; 95% CI,1.63-4.13), bronchiectasis (OR=1.40; 95% CI,1.20-1.61), and duodenal ulcer (OR=1.30; 95% CI,1.15-1.45) (**Fig. 2a** and **Table S2**). Four additional associations reached significance at a relaxed FDR threshold of 10%: COPD (OR=1.17; 95% CI 1.07-1.27), cholelithiasis (OR=1.13; 95% CI 1.06-1.22), male infertility (OR=2.10; 95% CI,1.40-3.15), and other prostate disorders (OR=1.39; 95% CI 1.15-1.67) (**Table S2**).

**Figure 2.**
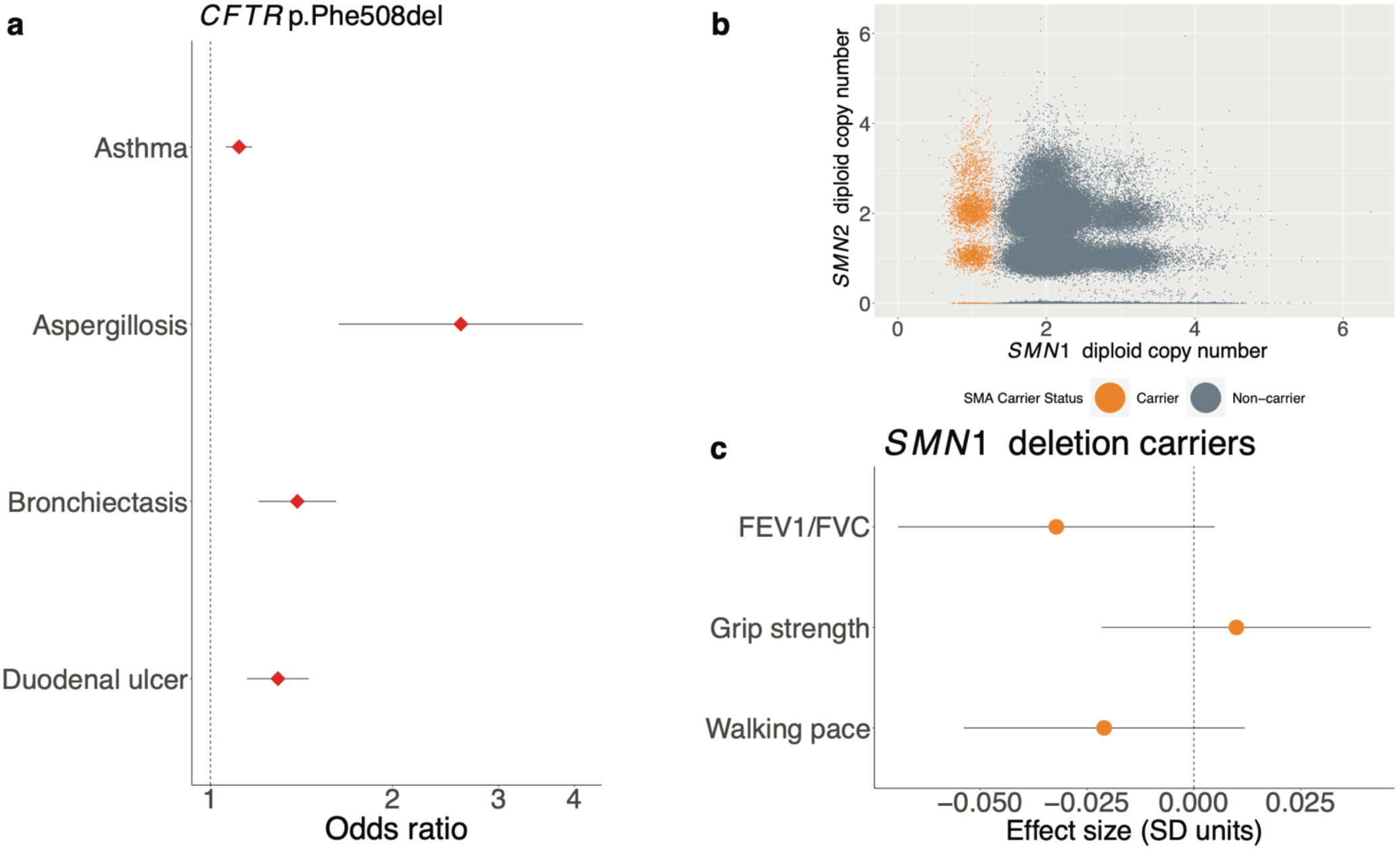
Cystic fibrosis carriers show mitigated phenotypes, but spinal muscular atrophy carriers do not. (**a**) Carriers with the *CFTR* Phe508del inframe deletion exhibit increased risk of several mitigated disease phenotypes (data points, odds ratios; error bars, 95% CIs). (**b**) Genotyping for SMA carrier status using exome sequencing data from the UK Biobank. *SMN1* and *SMN2* copy numbers were estimated based on sequencing read depth, and SMA carriers (orange) were identified as those individuals estimated to have one functional copy of *SMN1* (with a deletion allele on the homologous chromosome). (**c**) *SMN1* deletion carriers did not display evidence of changes in any of three traits related to neuromuscular function (data points, mean values in units of standard deviations; error bars, 95% CIs).

We also tested carriers of the next most common cystic fibrosis mutation, *CFTR* Arg117His (MAF=0.2% in UK Biobank) but concluded that power was insufficient (**Table S2**).

The odds ratios we calculated for carriers of Phe508del, while significant, were much smaller than those recently reported in an analysis of CF carriers ascertained from a database of insurance claims from individuals who had been tested for carrier status^4^ (**Fig. S1**). Furthermore, several reported associations did not replicate in our analysis of UK Biobank. For example, whereas the claims analysis showed a strong association between carrier status and short stature^4^, we did not observe an association between Phe508del carrier status and height in UK Biobank despite ample power (effect size = -0.000 ± 0.006 SD). The odds ratios we computed were more consistent with those reported by Çolak et al. (2020) using Phe508del genotyping in the Copenhagen General Population Study^5^. These results underscore the importance of understanding issues of ascertainment bias when studying penetrance in population studies^51–53^.

In contrast to CF, potential phenotypes of SMA carriers have not (to our knowledge) previously been explored, in part because of the difficulty of genotyping SMA carrier mutations, most of which arise from structural variation at the *SMN1*–*SMN2* locus^29,54^. SMA is usually caused by loss-of-function mutations in both copies of the *SMN1* gene, with disease severity then determined by the number of functional copies of the paralogous *SMN2* gene. The availability of whole-exome sequencing (WES) data for *N*∼200K UK Biobank participants^19^ enabled us to estimate the number of functional copies of *SMN1* and *SMN2* in each sequenced sample from WES depth-of-coverage (**Fig. 2b**). We ascertained 3,462 SMA carriers (i.e., individuals likely to carry only one functional copy of *SMN1*) in this way from the set whole-exome sequenced individuals of European-ancestry (*N*=187,720). Interestingly, we found no significant associations between SMA carrier status and potential manifestations of muscle weakness – walking speed, grip strength, and FEV1/FVC ratio (**Fig. 2c**) – even when stratifying for *SMN2* copy number (**Table S3**). These results suggest that SMA is a truly recessive disease in which muscle weakness phenotypes only manifest in individuals who carry two *SMN1* alleles inactivated by loss-of-function variants.

### Testing a model of modified penetrance

In all the instances in which we observed mitigated phenotypes in carriers of recessive disease variants, the associated phenotypes exhibited variable penetrance in heterozygotes. We therefore sought to explore the possible molecular mechanisms underlying this incomplete penetrance.

Castel *et al*. (2018) previously proposed a model of modified penetrance in which the haplotype arrangement of loss-of-function and expression-modifying variants in an individual might affect overall phenotype (**Fig. 3a**)^23^. In this model, the phenotypic impact of a deleterious variant inactivating one copy of a gene is mediated by the amount of expression of the functional copy (on the opposite haplotype), such that a common *cis*-eQTL influencing expression of the functional allele can influence the severity of the phenotype. Explicitly, if the *cis*-eQTL increases expression of the functional, wildtype protein, this could partially ameliorate the loss of the other copy; in contrast, if the *cis*-eQTL decreases expression of the functional copy, one might expect the carrier to have a more severe phenotype.

**Figure 3.**
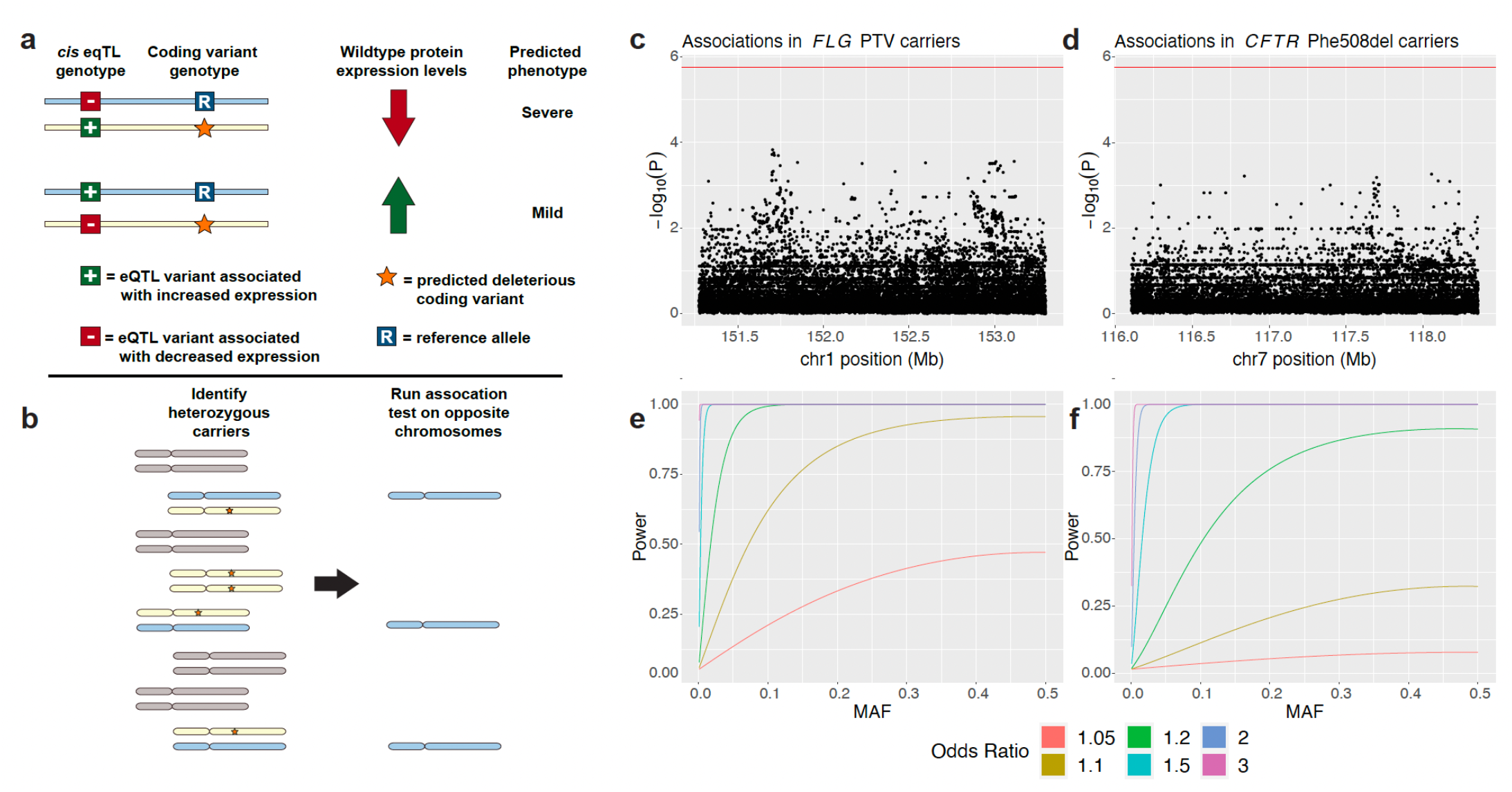
Common variants on haplotypes opposite recessive alleles in carriers of two Mendelian diseases do not appear to modify penetrance of carrier phenotypes. (**a**) The “modified penetrance” model of Castel *et al*. (2018) posits that *cis*-eQTLs can increase or decrease severity of a deleterious variant by modulating the quantity of functional protein produced by the opposite haplotype. (**b**) To test this hypothesis, we examined opposite haplotypes (blue chromosomes) in heterozygous carriers of recessive disease variants (orange star on yellow chromosome). We tested variants carried on these opposite haplotypes for association with mitigated phenotypes observed in carriers. (**c**,**d**) Manhattan plots showing association test results for variants on the opposite haplotype of deleterious variants in *FLG* and *CFTR* with asthma (the phenotype most strongly associated with carrier status in each case). No association reached Bonferroni significance (red line). (**e**,**f**) Power analyses for the tests conducted in **c**,**d** indicate that these tests were well-powered to detect common variant effects with odds ratios >1.2.

To explore this hypothesis, we considered two genes, *FLG* and *CFTR*, in which variants known to cause both recessive disease and produce mitigated phenotypes in carriers are sufficiently common to power analysis. Loss-of-function variants in *FLG* are known to cause ichthyosis vulgaris in homozygotes or compound heterozygotes, and carrier status has been associated with asthma and atopic dermatitis^55,56^. In UK Biobank, 10.3% of participants carried a loss-of-function variant in *FLG* that was associated with asthma or atopic dermatitis in heterozygotes. As discussed in the previous section, mutations in *CFTR* are responsible for cystic fibrosis, as well as several mitigated phenotypes in carriers. Approximately 3.1% of individuals in the UK Biobank are carriers for the Phe508del mutation in *CFTR* that we considered for this analysis.

To determine whether variants on the opposite (putatively functional) haplotype in carriers might affect their susceptibility to mitigated phenotypes, we restricted our analysis just to carriers of these deleterious variants (**Fig. 3b**). For each nearby variant at each locus, we then ran an association test between opposite-haplotype genotypes and mitigated phenotypes. No tested variant at either locus significantly associated with phenotype (**Fig. 3c**). Given that we were well-powered to detect common variant associations with an odds ratio >1.2 in both scenarios (**Fig. 3d**), these results suggest that the modified penetrance model is unlikely to underlie incomplete penetrance of these carrier phenotypes.

## Discussion

Our results demonstrate that for a wide range of Mendelian diseases, variants traditionally considered to be recessive can cause milder phenotypes in heterozygous carriers. We also observed that entirely recessive effects do exist: heterozygous carriers for spinal muscular atrophy exhibited no evidence of even a subtle effect on phenotypes related to muscle strength. These observations suggest a spectrum of recessiveness that is now becoming visible in very large population cohorts.

Our study did have several limitations. First, even with the large sample size provided by exome sequencing in UK Biobank, we still lacked power to assess potential effects of many very rare variants that are known to cause Mendelian recessive diseases. Second, our examination of potential interactions between variants on opposite haplotypes was even more power-constrained, such that we could only assess this model for two diseases involving common variants. Third, the effects we estimated are likely to be influenced by the “healthy volunteer” ascertainment bias observed in analyses of population biobank cohorts^22^.

As even larger, well-phenotyped cohorts with whole-exome or whole-genome sequencing become available, our ability to determine the extent of mild carrier phenotypes will increase. More comprehensive phenome-wide and genome-wide studies will allow for an assessment of how common the phenomenon of incomplete recessivity is amongst severe Mendelian diseases and the spectrum of phenotypes that can manifest. Moreover, the higher power afforded by extremely large studies will also enable more extensive exploration of potential interactions between variants that could help to explain incomplete penetrance and shed light on the molecular mechanisms that underlie mitigated phenotypes.

## Data Availability

Access to the UKB Resource is available by application (http://www.ukbiobank.ac.uk/).

## Supplemental data

include one figure and three tables.

## Acknowledgments

We thank A. Gusev, A. Price and S. Sunyaev for helpful discussions. This research was conducted using the UK Biobank Resource under application no. 10438. A.R.B. was supported by US NIH grant T32 HG229516 and fellowship F31 HL154537. M.L.A.H. was supported by US NIH fellowship F32 HL160061. M.A.S. was supported by the MIT John W. Jarve (1978) Seed Fund for Science Innovation and US NIH fellowship F31 MH124393. R.E.M. was supported by US NIH grant K25 HL150334 and NSF grant DMS-1939015. P.-R.L. was supported by US NIH grant DP2 ES030554, a Burroughs Wellcome Fund Career Award at the Scientific Interfaces, the Next Generation Fund at the Broad Institute of MIT and Harvard, and a Sloan Research Fellowship. The funders had no role in study design, data collection and analysis, decision to publish or preparation of the manuscript. Computational analyses were performed on the O2 High Performance Compute Cluster, supported by the Research Computing Group, at Harvard Medical School (http://rc.hms.harvard.edu).

## Data and code availability

Access to the UK Biobank Resource is available by application (http://www.ukbiobank.ac.uk/). BOLT-LMM (v2.3.4) is available at https://data.broadinstitute.org/alkesgroup/BOLT-LMM/. mosdepth (v0.3.1) is available at https://github.com/brentp/mosdepth. Minimac4 (v.1.0.1) is available at https://genome.sph.umich.edu/wiki/Minimac4. plink (v1.9) is available from https://www.cog-genomics.org/plink1.9/.

## Web resources

ClinVar, https://www.ncbi.nlm.nih.gov/clinvar/

OMIM, https://omim.org/

