## Supplemental Figure 1 for "A spectrum of recessiveness among Mendelian disease variants in UK Biobank"

**Figure S1. Comparison between odds ratios for mitigated disease phenotypes in CF carriers in UK Biobank versus results from Miller *et al*. (2020)**. Results for this analysis of UK Biobank are shown as red circles; results from Miller *et al*. are shown as black triangles (data points, odds ratio; error bars, 95% CIs).

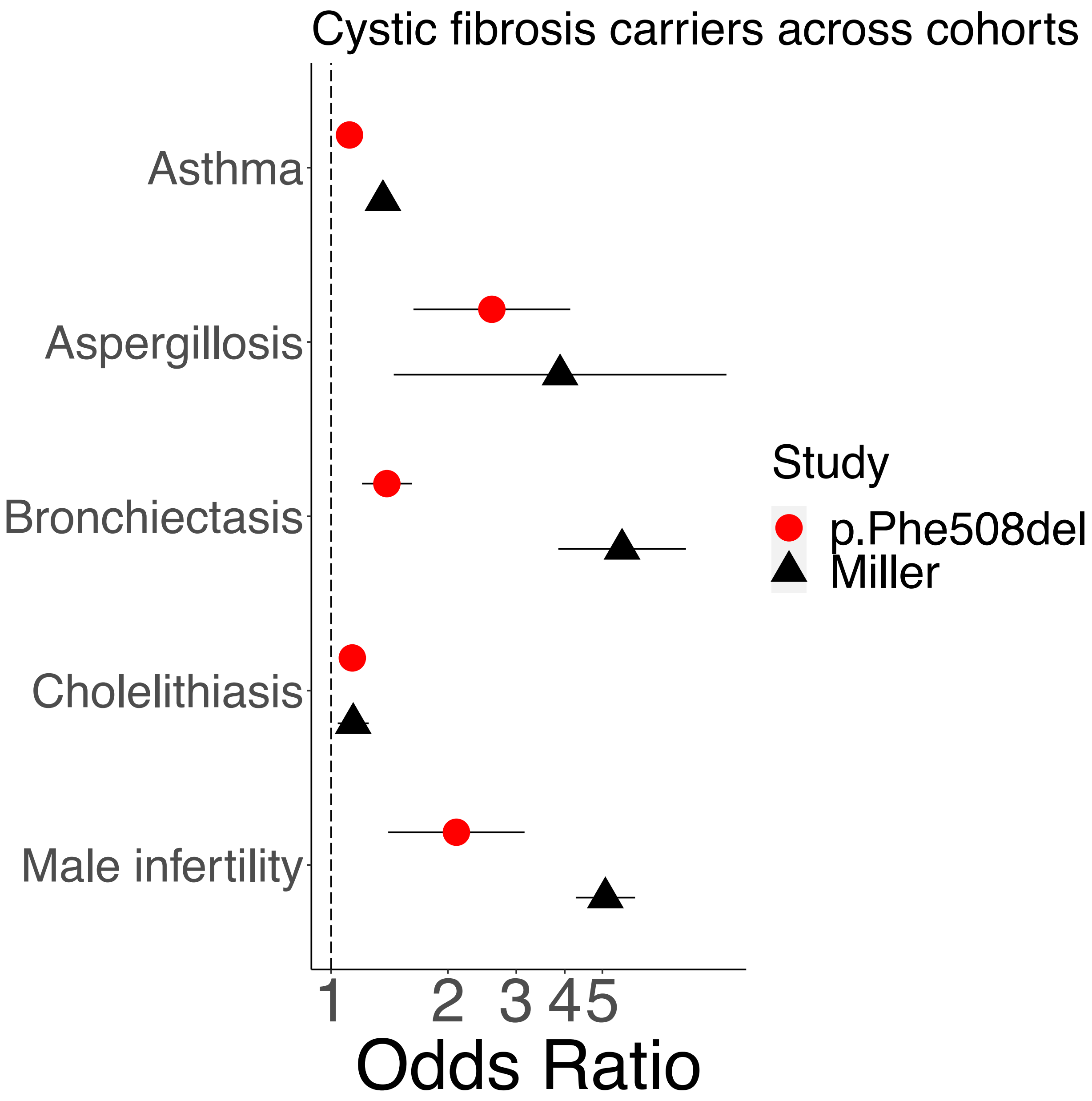
